# Finding Rare Disease Patients in EHR Databases via Lightly-Supervised Learning

**DOI:** 10.1101/2020.07.06.20147322

**Authors:** Rich Colbaugh, Kristin Glass

## Abstract

There is considerable interest in developing computational models capable of detecting rare disease patients in population-scale databases such as electronic health records (EHRs). Deriving these models is challenging for several reasons, perhaps the most daunting being the limited number of already-diagnosed, ‘labeled’ patients from which to learn. We overcome this obstacle with a novel lightly-supervised algorithm that leverages unlabeled and/or unreliably-labeled patient data – which is typically plentiful – to facilitate model induction. Importantly, we prove the algorithm is *safe:* adding unlabeled/unreliably-labeled data to the learning procedure produces models which are usually more accurate, and guaranteed never to be less accurate, than models learned from reliably-labeled data alone. The proposed method is shown to substantially outperform state-of-the-art models in patient-finding experiments involving two different rare diseases and a country-scale EHR database. Additionally, we demonstrate feasibility of transforming high-performance models generated through light supervision into simpler models which, while still accurate, are readily-interpretable by non-experts.

## 1. Introduction

Taken together, rare diseases affect more than 350M individuals worldwide, and many are chronic, progressive, degenerative, and life-threatening. They also tend to be unfamiliar and challenging to recognize, requiring an average of five years and seven physicians to be assigned an accurate diagnosis (if diagnosed at all) [1-4]. Difficulty finding patients who are suffering from a given rare disease is a major impediment to effective diagnosis and treatment, and is an obstacle to achieving many other clinical and research goals (e.g. cohort recruitment, disease characterization, resource allocation). Because the prevalence of each rare disease is low, practically-useful patient-finding must be pursued using population-scale databases such as electronic health records (EHRs) or Web activity logs, and therefore must be realized via computational modeling [5-9]. While it is possible to derive the models through consultation with experts, this strategy can be problematic for several reasons, including the incomplete nature of experts’ understanding, the complexity of disease processes, and cognitive biases associated with human decision-making [10-12,2].

These considerations motivate an interest in learning computational models for patient discovery directly from data [e.g. 13], and in particular by applying predictive modeling to EHRs [7-9,14-16]. This scheme faces difficulties of its own, perhaps the most crucial being that, with rare diseases, there are only a limited number of already-diagnosed patients available for model training. EHR databases possess additional properties which must be addressed for successful data-driven modeling, including: i.) their large-scale and high-dimensionality, ii.) the unreliable, incomplete, and sparse character of the data, iii.) the fact that patient records combine a myriad of structured (e.g. hierarchically-organized diagnostic codes) and unstructured (e.g. free-text notes) information, the vast majority of which is irrelevant to the prediction task at hand. See [7-9,14-16] for further discussion of these issues.

Given a target disease of interest, perhaps the most natural way to build patient-finding models is through *supervised learning*. Standard supervised learning in this context entails: i.) labeling the EHRs of already-diagnosed patients as being positive for the disease, ii.) adding these records to those of some unaffected patients, who form the negative class, to assemble a training dataset, and iii.) applying a validated machine learning algorithm to this training data and in this way acquire a model capable of predicting the disease status of new unseen patients [17]. Unfortunately, if the target disease is rare, then there are invariably too few patients with confirmed diagnoses to support construction of generalizable models using standard supervised techniques [17,8].

Perhaps surprisingly, recent work has shown it is sometimes possible to learn accurate predictive models by combining limited labeled data with additional, less directly-informative, sources of supervision, and exploiting this ‘light’ supervision to guide the modeling process [18]. For instance, training examples with unreliable or missing labels can represent valuable sources of light supervision if they are appropriately leveraged [8,18-21]. Observe that this *lightly-supervised* approach to learning appears to represent a promising alternative in the present setting because EHR databases typically contain [8]: i.) an abundant supply of patient records which are *unlabeled* relative to the target disease, and ii.) patients whose records document some evidence of positive target-disease status but who do not have confirmed diagnoses, and who therefore may be assigned tentative, *noisy labels*.

We have developed a novel learning methodology which enables accurate patient-finding models to be induced from the imperfect real-world data encountered when investigating rare diseases. The approach enjoys two key attributes. First, learning requires only the light supervision provided by a few reliably-labeled patients, some patients with unreliable/noisy labels, and a cohort of patients who are unlabeled. Second, learning is provably *safe:* adding unlabeled and noisily-labeled data to the learning procedure produces models which are usually more accurate, and never less accurate, than models obtained with the reliably-labeled data alone. The safety guarantee is important. While supervision from noisy/incomplete data is intended to improve learning performance when reliably-labeled examples are limited, existing algorithms may not actually achieve this aim. Indeed, incorporating less directly-informative data often yields models that perform worse than those found using the (limited) reliable data alone [18-21].

In what follows, we describe and validate this new approach to rare-disease patient-finding. More specifically, this paper:

- introduces a new algorithm for learning prediction models from the light supervision delivered by a few reliably-labeled patients, some patients with noisy labels, and a set of patients who are unlabeled relative to the (rare) disease of interest;
- proves the algorithm is *safe:* the addition of unlabeled/noisily-labeled examples to a small training set of reliably-labeled data, to compensate for its limited size, yields models which are usually more accurate, and never less accurate, than corresponding models induced from the original training set;
- demonstrates that the proposed strategy outperforms state-of-the-art models for the task of detecting two different rare diseases in a country-scale database of 3.2M EHRs;
- shows the feasibility of transforming learned high-performance patient-finding models – which consist of large ensembles of complex prediction models – into simpler, but still accurate, models that are interpretable by non-experts.

## 2. Safe Lightly-Supervised Learning

### A. Problem Formulation

Consider a scenario in which we are given a target rare disease (TD) and access to a database of de-identified EHRs, and wish to learn a prediction model capable of reliably detecting (undiagnosed) patients with the disease in the database. Ideally, learning should be feasible even if only a few patients known to have the disease are available for model training. To focus on the most widely-applicable setting, it is assumed the EHRs document patient encounters with general practitioners (GPs) (rather than, say, specialists) and contain standard information: basic demographic data, diagnosis codes, medication codes, free-text clinical notes, and limited lab test results (because lab test results are frequently not reported in GP EHRs). We refer to such a database as GP-EHR-DB and represent each patient in it with a vector of measurement values (e.g. age, blood glucose level) and counts of the various ‘tokens’ (e.g. diagnostic codes, words); see below for additional details.

As indicated above, data-driven patient-finding through supervised learning consists of three basic steps [17]:

1. model each patient as (**x**,y), where feature vector **x** = [x_1_, …, x_d_]^T^∈X encodes information in the patient’s EHR [e.g. 14] and label y∈{0,1} denotes whether **x** has TD;
2. assemble a set of patient EHRs L = {(**x**_1_, y_1_), …, (**x**_M_, y_M_)}, where the disease status of each of these patients is known and thus labels y_i_ are reliable (conventionally y_i_ = 1 if **x**_i_ has TD);
3. use labeled dataset L to learn model **f**_L_: X → [0,1] which predicts the probability p(y = 1 | **x**) that a given patient **x** has TD [17].

A challenge with predictive modeling of rare diseases is that the number of training examples M is invariably small and the ‘light’ supervision supplied by L is insufficient to support effective learning [17,8]. In fact, dataset L may be empty (M = 0). It turns out this situation also can be analyzed with the proposed methodology by specifying a ‘prior’ for the probability that patients in GP-EHR-DB have TD (e.g. based on prevalence estimates, perhaps stratified by patient phenotype). This extension is illustrated below with one of the target rare diseases investigated in this paper.

Because (standard) supervised learning is not well-suited to the task of finding rare-disease patients, we introduce a novel strategy which exploits limited supervision to learn high-performance models, that is, models capable of accurately predicting the TD-status of all patients in GP-EHR-DB. This lightly-supervised learning (LSL) strategy integrates three sources of supervision (dataset L is listed again for completeness):

- *reliably-labeled patients* L = {(**x**_1_, y_1_), …, (**x**_M_, y_M_)}, where labels y∈{0,1} are accurate but number of patients M is too small to facilitate generalizable learning;
- *noisily-labeled patients* N = {(**x**_M+1_, y′_M+1_), …, (**x**_M+N_, y′_M+N_)}, where some patients in N are expected to be mislabeled, that is, y′ ≠ y with (unknown) probability p_noise_;
- *unlabeled patients* Tar = {**x**_M+N+1_, …, **x**_M+N+T_}; these are the patients for whom TD-status is to be predicted (i.e. the labels for these patients are unknown).

With these definitions in place, the safe LSL problem can be stated: given a target disease TD and sets of reliably-labeled, noisily-labeled, and unlabeled patient EHRs L, N, and Tar, respectively, learn a patient-finding model that normally outperforms **f**_L_, learned using L alone, and is guaranteed to never perform worse than **f**_L_.

### B. Overview of Learning Method

The proposed scheme for learning rare-disease patient-finding models in a safe lightly-supervised manner consists of three main steps. First, labeled dataset L is used to induce prediction model **f**_L_: X → p_0_, where p_0_ is the ‘baseline’ predicted probability patient **x**∈Tar has label y = 1 (i.e., has TD). Applying **f**_L_ to unlabeled dataset Tar yields a vector of predictions **p**_0_ = [p_0(M+N+1)_, …, p_0(M+N+T)_]^T^, one for each of the T patients in Tar.

However, if the number of labeled examples M is small, the predictions **p**_0_ may not be accurate. This observation motivates the second step in the learning strategy: construct an ensemble of additional predictions which can be exploited to improve upon baseline **p**_0_. Because it is advantageous for the ensemble to be diverse, we generate the predictions which comprise it using two different techniques:

- *prediction refinement* of **p**_0_ guided by the underlying structure of the data, as revealed through unsupervised clustering of unlabeled patient dataset Tar;
- *noise-robust learning* informed by both unreliably-labeled dataset N and the underlying structure of patient data.

While predictions made with these two methods are typically informative, attempting to use them to improve predictions **p**_0_ may actually *decrease* accuracy, rather than increase it. Thus the third step in the learning methodology is to *safely* combine **p**_0_ with the ensemble predictions, that is, to integrate this information to form final predictions which are ordinarily better, and never worse, than **p**_0_.

The first step in the prediction process – inducing model **f**_L_ from labeled dataset L – is straightforward to complete using any algorithm which allows statistically-efficient supervised learning. Here we adopt a random forests (RF) model for **f**_L_, with 1000 trees and default hyperparameter values [17] (varying tree number between 200 and 2000 had modest effect).

The second and third steps in the SLSL procedure are more involved, warranting the separate descriptions provided in the next two sections.

### C. Ensembles of Predictions

This section recalls two analytic methods, which we first introduced in [22] in a different context, for building ensembles of predictions.

#### Prediction refinement

As indicated above, labeled dataset L can be used to induce prediction model **f**_L_: X → p_0_, where p_0_ is the predicted probability that patient **x**∈Tar has label y = 1. Applying **f**_L_ to unlabeled dataset Tar yields vector of baseline predictions **p**_0_ = [p_0(M+N+1)_, …, p_0(M+N+T)_]^T^, one for each of the T patients in Tar. **p**_0_ can be used to create an ensemble of diverse predictions by leveraging unlabeled patients in Tar to create refinements of this baseline [22]. The assumption behind the refinement strategy is that if patients **x**_k_ and **x**_l_ are ‘similar’ then they should possess similar labels.

More precisely, patient similarity is used to refine **p**_0_ to **p**′ = [p′_M+N+1_, …, p′_M+N+T_]^T^ with p′_i_ the predicted probability instance **x**_i_ has label y_i_ = 1. Toward this end, let La = I − C denote a Laplacian matrix, where I is the identity matrix and C a similarity matrix computed via ensemble clustering on Tar [17]. C is constructed so that element C_kl_ is equal to the number of times patients **x**_k_ and **x**_l_ are assigned to the same cluster by members of the ensemble. (Varying the number of clusters per ensemble member and number of ensemble members has little impact on performance. Reasonable choices for these hyperparameters are 2-20 clusters and 5-20 ensemble members.) Predictions **p**′ are formed by balancing the goals of achieving within-cluster label similarity and maintaining agreement with the baseline predictions **p**_0_:

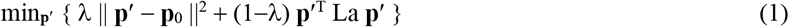

subject to the constraint that each p_i_∈[0,1], where λ∈[0,1] reflects the relative expected predictive value of labeled and unlabeled data.

The optimization (1) can be accomplished by iterating the following formula over j until convergence (which is assured [23,24]):

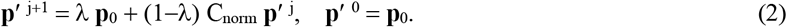

In (2), C_norm_ is the normalized version of C obtain by expressing C as a symmetric probability matrix. This solution is computationally efficient, allowing population-scale problems to be investigated.

Finally, an ensemble of R1 prediction refinements {**p**′^1^, …, **p**′^R1^} can be assembled by varying λ and/or defining different Laplacian matrices La through the use of distinct clusterings of Tar (see the experiments below).

#### Noise-robust learning

Noisily-labeled dataset N can also be exploited to model the probability that patient **x**∈Tar has label y = 1. However, effectively using noisy labels requires that learning be robust to mislabeling. We achieve this robustness using a scheme similar to the one summarized in the preceding subsection (see also [8]). More precisely, ‘preliminary’ predictions are made for patients in Tar using a model learned from dataset N, and these predictions are then improved using information which is *independent* of the noisy labels.

RFs are known to be robust to label noise [17,25], especially if built from reasonably shallow trees (larger leaf nodes can ‘average out’ label noise). We therefore learn ‘robustified’ RFs by modifying the algorithm to penalize excessive tree-depth [25]. In this way, noisily-labeled dataset N is used to induce prediction model **f**_N_: X → p_0_′, where p_0_′ is the predicted probability that patient **x**∈Tar has label y = 1. Applying **f**_N_ to unlabeled dataset Tar gives preliminary predictions **p**_0_′ = [p_0_′_(M+N+1)_, …, p_0_′_(M+N+T)_]^T^ for the patients in Tar.

While preliminary predictions **p**_0_′ are expected to be useful, it can be helpful to mitigate the influence of label noise on **p**_0_′ with the procedure (2), rewritten here for convenience of reference (see also [25]):

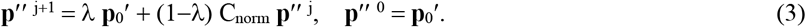

This computation yields **p**′′ = [p′′_M+N+1_, …, p′′_M+N+T_]^T^, where p′′_i_ is the predicted probability **x**_i_∈Tar has label y_i_ = 1. As before, an ensemble of predictions {**p**′′^1^, …, **p**′′^R2^} can be acquired by varying λ and/or producing a set of Laplacian matrices using distinct clusterings of Tar (see the experiments below).

### D. Algorithm SLSL

We now show how the ensemble of predictions Π = {**p**′^1^, …, **p**′^R1^, **p**′′^1^, …, **p**′′^R2^} can be leveraged to improve baseline predictions **p**_0_ made with model **f**_L_ learned from (small) labeled dataset L. Importantly, the proposed methodology is *safe:* the final predictions **p** are guaranteed to be at least as accurate as **p**_0_.

It is natural to approach safe LSL (SLSL), in which assurances are sought for prediction quality in the presence of uncertainty, as an adversarial learning problem [26]. Toward this end, let **y** = [y_M+N+1_, …, y_M+N+T_]^T^ denote the true (hidden) labels for patients Tar = {**x**_M+N+1_, …, **x**_M+N+T_}, and define **α** = [α_1_, …, α_R1+R2_]^T^ with α_i_ ≥ 0 and ∑_i_α_i_ = 1. **α** can be employed to specify various convex combinations of the predictions Π via **p**_c_ = Π**α**, and in this way model an ‘adversary’ within the learning process. Informally, suppose the true labels **y** satisfy **y** = Π**α**_true_ for some unknown **α**_true_. Then our objective is to form predictions **p** which are close to Π**α**_true_ despite our uncertainty about **α**_true_. If an adversary is introduced whose goal is to choose **α** to make prediction difficult, this provides a framework within which to seek predictions **p** that are ‘good’ even in worst-case, that is, when **α**_true_ is close to the adversarially-chosen value for **α**.

These considerations can be formalized by stating a game theory-inspired algorithm [25-27,21] for computing predictions **p** which improve upon **p**_0_ even in worst-case.

#### Algorithm SLSL

1. Learn model **f**_L_ from dataset L and use it to form baseline predictions **p**_0_ = [p_0(M+N+1)_, …, p_0(M+N+T)_]^T^ for unlabeled patient dataset Tar = {**x**_M+N+1_, …, **x**_M+N+T_}.
2. Learn model **f**_N_ from dataset N and use it to form preliminary predictions **p**_0_′ = [p_0_′_(M+N+1)_, …, p_0_′_(M+N+T)_]^T^ for unlabeled patient dataset Tar.
3. Assemble an ensemble of predictions Π = [**p**′^1^, …, **p**′^R1^, **p**′′^1^, …, **p**′′^R2^] by: i.) refining **p**_0_ → {**p**′^1^, …, **p**′^R1^} via (2) and ii.) adapting **p**_0_′ → {**p**′′^1^, …, **p**′′^R2^} using (3) (see Section 2.C for details).
4. Compute final predictions **p** by solving the following optimization problem [25-27,21]

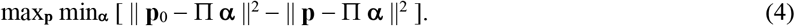

It is seen that (4) quantifies the adversarial setting summarized above: we select predictions **p** to maximally improve upon **p**_0_ in worst-case, that is, when true labels **y** = Π**α**′ with **α**′ chosen by the adversary to minimize any such improvement in prediction accuracy.

The safety and effectiveness of Algorithm SLSL is established in

**Theorem SLSL:** Assume there exists (unknown) **α**_true_ such that the true labels **y** = Π **α**_true_. Then the optimal solution (**p**\*, **α**\*) to (4) ensures i.) ‖ **p**\*− **y** ‖ ≤ ‖ **p**_0_ − **y** ‖ and ii.) **p**\* achieves maximum worst-case performance gain over **p**_0_.

**Proof:** It is easy to check that objective function

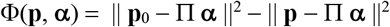

satisfies the conditions of the minimax theorem [27], so that

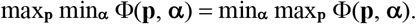

This, in turn, implies

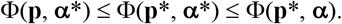

Setting **p** = **p**_0_ and **α** = **α**_true_ in this chain of inequalities gives

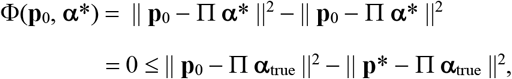

implying ‖ **p**\* − **y** ‖ ≤ ‖ **p**_0_ − **y** ‖, which is claim i. Claim ii. follows from this and the optimality of **p**\*. ▄

Thus predictions **p**\* = [p*_M+N+1_, …, p*_M+N+T_]^T^ for the TD-status of patients Tar = {**x**_M+N+1_, …, **x**_M+N+T_} are always at least as close to the true labels **y** as baseline predictions **p**_0_, and improve upon **p**_0_ to the maximum extent possible in worst-case (when **y** = Π**α**\*).

In addition to being provably-safe, Algorithm SLSL is efficient to execute. For instance, it is easy to show that optimization problem (4) is convex, implying that optimal predictions **p**\* can be computed via standard convex quadratic programming [28].

## 3. Patient-Finding Experiments

We now describe a study in which Algorithm SLSL is deployed to find patients suffering from two very rare, difficult-to-diagnose TDs in a large GP-EHR-DB. The investigation:

- performs patient-finding for two quite different TDs in settings in which reliable labels are extremely limited (2 and 0 patients with confirmed diagnoses for the two TDs);
- illustrates construction of EHR training datasets in this challenging scenario;
- demonstrates that models learned with Algorithm SLSL outperform state-of-the-art benchmark models and enable high-precision detection of TD patients in a country-scale database of 3.2M EHRs;
- identifies those patient features which possess significant predictive power for each TD;
- illustrates the way results of chart review by TD specialists can be leveraged to obtain improved training labels and permit induction of even more accurate patient-finding models.

### A. Setup

The main goal of these experiments is to learn models which can accurately detect patients affected by a rare disease of interest using standard GP EHR data. In order to assess the generality of the approach, the experiments consider two TDs with quite distinct attributes:

- acute hepatic porphyria (AHP), an autosomal dominant disorder impacting heme synthesis and associated with serious and potentially life-threatening neurovisceral attacks; clinical presentation is heterogeneous and can include severe abdominal pain, nausea, vomiting, hypertension, tachycardia, and various neurological and psychiatric symptoms; prevalence is estimated to be around 5-20/1M [29,30];
- lipodystrophy (LD), a genetic/acquired disorder characterized by selective deficiencies of adipose tissue and associated with serious complications including diabetes mellitus, dyslipidemia, liver disease, heart disease, pancreatitis, and reproductive dysfunction; prevalence is estimated to be approximately 2-4/1M [31,32].

GPs may be unfamiliar with these diseases, and clinical manifestation of each is variable and can mimic many other more common diseases. Thus these TDs are often repeatedly misdiagnosed or not detected at all. Difficulty identifying patients leads to treatment delays, negatively impacts patient health, and slows progress in clinical research [1-4], highlighting the real-world relevance of this set of experiments.

Experiments were conducted using a database of 3.2M de-identified EHRs arising from patient encounters with GPs in a European country [33]; we refer to this database as GP-EHR-DB-NL. The information contained in GP-EHR-DB-NL includes basic demographics data, ICPC diagnosis and symptom codes (http://www.who.int/classifications/icd/adaptations/icpc2/), ATC drug codes (https://www.whocc.no/atc_ddd_index), and limited free text clinical notes and lab results. GP-EHR-DB-NL is representative of other de-identified GP EHR datasets and the challenges faced when analyzing them. For instance, clinical notes and lab test records are typically brief and incomplete, reflecting privacy concerns and the nature of GP visits.

Patients are represented as pairs (**x**, y), where feature vector **x** encodes a patient’s EHR information in the usual way. Briefly, **x** is composed of year-of-birth, gender, counts for all ICPC and ATC codes, numerical values for lab test results and clinical measurements, and a ‘bag-of-words’ model for free-text clinical notes (yielding high-dimensional patient vectors). Further details on this processing step can be found in [e.g. 14,8]. The label y designates TD-status, with y = 1 if the patient has the TD in question and y = 0 if not.

Training datasets were assembled in the following manner. For each TD, the *positive-class* examples consist of patients with confirmed diagnoses (2 for AHP, 0 for LD) together with a number of (noisily-labeled) patients who *may* have TD (i.e., their EHRs contain some evidence of TD). The *negative class* training set was gathered by randomly-sampling EHRs of i.) patients diagnosed with diseases having clinical presentation similar to TD (mimics), ii.) patients matched to positive-class examples for age, gender, and date of earliest record. The criteria employed for identifying positive-class patients were defined through consultation with domain experts [8]. To avoid data leakage, only information entered into an EHR prior to diagnosis of the pertinent condition (TD or mimic disorder) was retained for analysis. Similarly, all patient EHRs were edited to remove mentions of terms used in the dataset collection procedure.

Attributes of the training datasets are listed in Table 1. Notice that each TD has low prevalence, there are few/no already-diagnosed patients in GP-EHR-DB-NL, and patients are described by thousands of features. As mentioned, the noise and high-dimensionality of the dataset are key obstacles to learning accurate, generalizable models for rare disease patient-finding.

**Table 1.** Training datasets.

| Target Disease (TD) | Estimated Prevalence | # Confirmed Diagnosed Patients | # Positive Class Patients: TD-Similar | # Negative Class Patients: Mimics | # Negative Class Patients: Matched | # Features |
| --- | --- | --- | --- | --- | --- | --- |
| AHP | 5-20/1M | 2 | 50 | 100 | 100 | 7469 |
| LD | 2-4/1M | 0 | 12 | 100 | 100 | 7469 |

In both the AHP and LD patient-finding experiments, Algorithm SLSL is implemented in four steps as specified in Section 2.D and detailed below.

1. Baseline prediction **p**_0_ is made with an RF model **f**_L_: X → p_0_ learned from dataset L (1000 trees and default hyperparameters; see Section 2.B and [17]).
2. Preliminary prediction **p**_0_′ is formed with a noise-robust RF **f**_N_: X → p_0_′ learned from dataset N (see Section 2.C and [8]).
3. Ensemble predictions Π = [**p**′^1^, **p**′^2^, **p**′^3^, **p**′′^1^, **p**′′^2^, **p**′′^3^] are made by: i.) refining **p**_0_ → {**p**′^1^, **p**′^2^, **p**′^3^} via (2) with three clusterings of Tar (k-means method [17] with k∈{2,4,7}) and ii.) adapting **p**_0_′ → {**p**′′^1^, **p**′′^2^, **p**′′^3^} using (3) with three clusterings of Tar (k-means, k∈{2,4,7}). Hyperparameter λ is set to λ=0.6 in iterative algorithms (2) and (3) (selected through nested cross-validation [17]).
4. Final prediction **p**\* is made by solving max_**p**_ min_**α**_ [‖ **p**_0_ − Π **α** ‖^2^ − ‖ **p** − Π **α** ‖^2^] using standard convex quadratic programming [28] (see Section 2.D).

Because intermediate and final predictions generated by Algorithm SLSL can be computed efficiently [23-25,28], patient-finding can be conducted using country-scale EHR databases.

To permit an objective, quantitative assessment of the performance of Algorithm SLSL, we compare the accuracy of its predictions with those of two benchmark models:

- ‘Anchor and learn algorithm’ [34,35]: a state-of-the-art technique for learning disease-state estimation models with very limited labeled data;
- ‘TD-training’ model: assume the TD training dataset is correctly-labeled and use all positive-class patients (i.e., the combination of already-diagnosed patients and patients for whom evidence exists of positive disease-status) as predictions for which patients have TD; this ‘model’ is similar to standard patient-finding methodologies, in which expert-derived code/keyword queries are employed to identify candidate patients [8]; its use here also allows the noise-level of training labels to be quantified.

We considered using standard supervised learning for this task (support vector machines, L1-regularized logistic regression [17]) but these models performed poorly in a pilot examination, likely because of the limited, noisy nature of the training data.

### B. Results

Algorithm SLSL is now applied to the task of finding undiagnosed TD patients in country-scale EHR database GP-EHR-DB-NL. Patient-finding is implemented in three steps:

1. Replace missing EHR fields with a ‘missing’ token [17].
2. Train Algorithm SLSL using the training dataset assembled for the TD under study (AHP or LD, see Table 1). Use the learned model to predict the probability that each patient in GP-EHR-DB-NL has the TD.
3. Rank-order patients by predicted probability of having TD and extract the Top 20 and Top CP candidates (with the threshold CP determined based upon natural breakpoints in the probabilities).

For each TD, we recruited appropriate specialists practicing in Europe to review the EHRs of the top CP candidates returned by our models. To increase reliability and objectivity of the reviews, the order of the patients was shuffled and 10 randomly-chosen patients were added to each list [8]. The specialists classified each candidate as ‘likely TD’, ‘possible TD’, ‘unlikely TD’, ‘highly unlikely TD’, ‘not TD’, or ‘unable to assess’. The results of the reviews are reported in Tables 2 and 3. Taken together, these results demonstrate that Algorithm SLSL is able to learn accurate patient-finding models from light-supervision provided by a few (two or zero) labeled examples together with noisily-labeled/unlabeled data. Indeed, the Top 20 Lists for AHP and LD achieve 90% precision and 70% precision, respectively, substantially outperforming state-of-the-art in this setting. To place these results in context, observe from current prevalence data (Table 1) that ∼0.001% of patients are expected to have AHP and ∼0.0003% of patients are expected to have LD.

**Table 2.** Results of patient-finding experiments: AHP.

|  | Total<br>Candidates (K) | Likely<br>AHP | Possible<br>AHP | Plausible<br>Candidates | Precision<br>@ K |
| --- | --- | --- | --- | --- | --- |
| Top 20 List | 20 | 14 | 4 | 18 | 90% |
| Top 42 List | 42 | 17 | 16 | 33 | 79% |
| AHP-Training List | 50 | 6 | 8 | 14 | 28% |
| Anchor and Learn | 33 | 6 | 6 | 12 | 36% |

**Table 3.** Results of patient-finding experiments: LD.

|  | Total<br>Candidates (K) | Likely<br>LD | Possible<br>LD | Plausible<br>Candidates | Precision<br>@ K |
| --- | --- | --- | --- | --- | --- |
| Top 20 List | 20 | 4 | 10 | 14 | 70% |
| Top 36 List | 36 | 4 | 15 | 19 | 53% |
| LD-Training List | 12 | 0 | 3 | 3 | 25% |
| Anchor and Learn | 15 | 1 | 4 | 5 | 33% |

In more detail, rank-ordering all patients in GP-EHR-DB-NL according to predicted probability of having AHP reveals the top 42 candidates have qualitatively higher AHP-probability than the rest (∼0.9). The AHP specialists reviewing the EHRs for these patients classified 17 as ‘likely AHP’ and an additional 16 as ‘possible AHP’, for a total of 33 plausible candidates (79% of the list). The top 20 list has even better precision, containing 14 ‘likely AHP’ patients and 4 ‘possible AHP’ patients, for a total of 18 plausible candidates (90% precision @ 20). In comparison, the Anchor and Learn model [34,35] nominates 33 potential AHP patients, of whom 12 are deemed to be plausible candidates (36%), while the 50-patient AHP-training list includes 14 plausible candidates (28%). The latter result shows that, as hypothesized, Algorithm SLSL can learn prediction models which are more accurate than the labels used for training. It can be seen that the models learned with Algorithm SLSL are also reasonably well-calibrated: the top 20 list has a greater ratio of ‘likely AHP’ to ‘possible AHP’ patients than the top 42 list. Se Table 2 for a summary of the experimental results for AHP.

By rank-ordering patients in GP-EHR-DB-NL according to predicted probability of having LD we find that the top 36 candidates have qualitatively higher LD-probability than the rest (∼0.8). This list of 36 candidates includes 4 patients that the specialist assessed to be ‘likely LD’ and 15 more deemed ‘possible LD’, for a total of 19 plausible candidates (53% of the list). The top 20 list has higher precision, containing the 4 ‘likely LD’ patients and 10 ‘possible LD’ patients, for a total of 14 plausible candidates (70% precision @ 20). As is the case with AHP, the Anchor and Learn method [34,35] provides much poorer performance, nominating 15 potential LD patients of whom 5 are deemed plausible candidates (33%). The 12-patient LD-training list includes 3 plausible candidates (25%), again indicating that Algorithm SLSL is capable of learning a prediction model which is more accurate than its training data. See Table 3 for a summary of the experimental results for LD.

Knowing which features captured in EHRs are predictive of a given rare disease is of value in various clinical applications. We assessed feature predictive power using both forward- and backward-stepwise analyses [17], and now highlight a few findings. The predictive power of structured v. unstructured features varies across the two TDs, with ICPC codes having significant predictive power for AHP, free text clinical notes being quite helpful for LD, and lab test results, though limited in these GP EHRs, being predictive for both AHP and LD. Proxies for temporal order of events are useful for distinguishing AHP from its mimics. Unusual combinations of otherwise common symptoms are also predictive (e.g. for LD, diabetes presenting in patients with normal or low body mass index (BMI)).

Fairly subtle characteristics of ‘patient journey’ can be informative. For example, longer sequences of unique ICPC codes, conjectured to reflect GP uncertainty/evolving understanding when confronted with a rare disorder, are predictive features for both AHP and LD, as are notes documenting GP behavior arising from uncertainty/unfamiliarity (e.g. multiple referrals or consultations). Finally, mental health-related clinical notes and medications (e.g. terms related to depression and anxiety, ATC codes/notes for anti-anxiety medications and anti-depressants) are distinguishing for both AHP and LD.

### C. Model Retraining with Specialist Labels

The experiments summarized in the preceding section demonstrate the accuracy and utility of patient-finding models learned by applying Algorithm SLSL to noisily-labeled/unlabeled training data. Interestingly, the chart review conducted by TD specialists in support of algorithm evaluation affords the opportunity to improve upon these models by leveraging TD-specialist appraisals. More specifically, we can replace the noisily-labeled positive-class training examples employed above with patient EHRs classified as ‘plausible TD’ by the specialists (i.e. ‘likely TD’ and ‘possible TD’ for each TD). Because the revised positive-class training sets are small, consisting of 33 and 19 patients for AHP and LD, respectively, Algorithm SLSL is still used for model-building (e.g. to permit unlabeled examples to be exploited).

This strategy does indeed generate very high-quality models. For instance, ten-fold cross-validation runs of the resulting models yield the following estimates for out-of-sample area under ROC curve (AUC):

- for AHP, AUC = 0.98;
- for LD, AUC = 0.96.

As another test of the new model, we rank-ordered the ‘top 42’ and ‘top 36’ lists for AHP and LD, respectively, according to the predicted TD-probabilities returned by the retrained patient-finding models. The reordered lists agree more closely with the specialists’ assessments.

## 4. Interpretable Models

### A. Basic Idea

The preceding theoretical (Section 2) and empirical (Section 3) discussions demonstrate that Algorithm SLSL offers a powerful and broadly-applicable approach to finding rare-disease patients in EHR databases. However, while the models learned by Algorithm SLSL are accurate, they are not *interpretable* by non-experts: each patient’s predicted TD-status is obtained by combining outputs of thousands of individual supervised and unsupervised models through convex quadratic programming. It would be desirable to use the algorithm as the basis for deriving detection models which are both accurate *and* interpretable [36-38], as this would enable clinicians to understand, and therefore trust, a model’s prediction process.

This section explores the feasibility of transforming the high-performance prediction models produced by Algorithm SLSL into accurate, interpretable models. The proposed technique accommodates key challenges associated with the clinical domain. Predictions concerning patient disease status are typically high-consequence and thus demand reliable, validated models. In addition, EHR data is characterized by missing, noisy, hierarchically-related features [39-41]. These problem requirements and attributes represent key obstacles for standard ways of achieving model interpretability. For instance, one common idea is to achieve interpretability by trading-off model performance for simplicity. However, the resulting models generally do not meet the performance demands of clinical tasks [37,38]. Alternatively, insights can be gained into the workings of a complex model by identifying which features are important to its predictions. Unfortunately, feature importance cannot be reliably-quantified for models that are designed to handle missing/noisy data or hierarchically-organized features [38-41].

We now present a simple method for learning accurate, interpretable patient-finding models which overcomes these challenges. The approach operationalizes a reasonable, mathematically-sound definition for model interpretability: a model is deemed *interpretable* if a user can take the model’s input data and parameters and, in reasonable time, execute the calculations necessary to make a prediction [36,37]. The basic algorithm is composed of three steps.

## Algorithm IM (Interpretable Modeling)

Given a target prediction task, interpretable models are learned in three steps.

1. Assemble a training set of patient EHRs (e.g. cohorts of patients known/suspected to either have or not-have the disease of interest).
2. Learn *full model* M_F_ which is capable of forming accurate predictions for new patients despite the practical difficulties associated with EHR-based modeling (e.g. missing feature values, unreliable patient labels, mix of structured and unstructured data) [8].
3. Abstract the full model M_F_ [42,43] to produce *interpretable decision tree model* M_IDT_ using one of the following methods:
  a. fit a decision tree to the *predictions* of M_F_ (rather than the patient labels appearing in the training dataset);
  b. abstract M_F_ using a slight extension of the logic-preserving (abstraction) algorithm put forth in [43].

A schematic of the procedure is depicted in Figure 1. In this feasibility study, only the model-abstraction 3a. is used; a more comprehensive investigation of model-abstraction and interpretable modeling, including the use of technique 3b., may be found in [38] (see also [43]).

**Figure 1.**
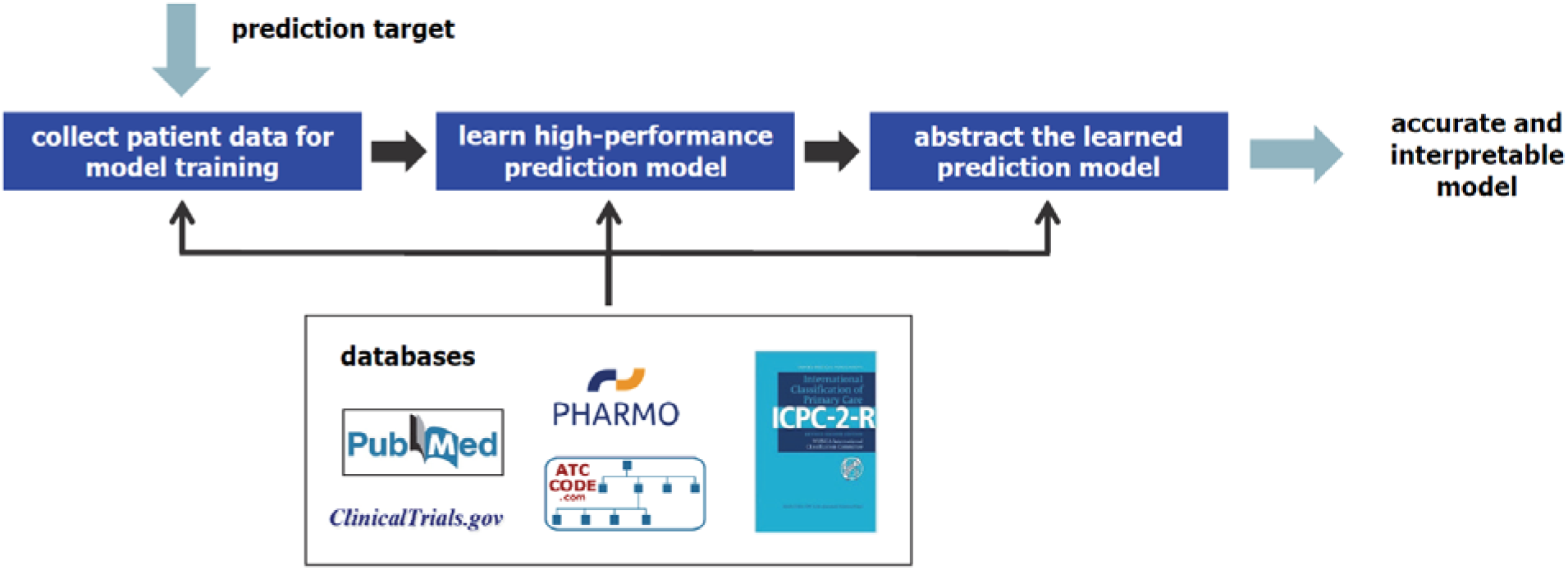
Schematic of interpretable modeling process.

While it is possible to apply the proposed interpretable-modeling procedure even with TDs that possess heterogeneous presentations spanning diverse medical domains, the resulting decision trees can be too complex to be readily-comprehensible. To address this issue we have designed an extension to the basic process, in which an ensemble of ‘specialist’ decision trees, rather than a single tree, is learned. For a given TD, the ensemble of specialists is induced by partitioning the full feature space into separate subspaces and learning one tree for each subspace [38]. The selection of feature subspaces is guided by the structure of medical domains, and consequently a learned ensemble tends to be composed of models which i.) align with medical specialties (e.g. cardiology, oncology), or ii.) correspond to aspects of the patient journey (e.g. by recognizing that GPs are imperfect ‘sensors’ and identifying feature subspaces that reflect clinical confusion). Interpretable specialist ensembles are illustrated in the next section and discussed in detail in [38].

### B. Experiments

The main goal of the experimental study is to evaluate the possibility of learning patient-finding models which are simultaneously accurate and interpretable. The task of interest is to detect undiagnosed AHP and LD patients in GP-EHR-DB-NL (3.2M patients). Algorithm IM is implemented as specified in Section 4.A. For each TD, model M_F_ is constructed by

- learning an initial model M_init_ using Algorithm SLSL;
- submitting patients predicted to have TD by M_init_ to disease specialists for chart review and revising the labels of the (positive-class) training dataset based upon this specialist assessment;
- training model M_F_ using the revised data;
- note: recall that models M_F_ are high-performance, achieving cross-validated accuracies of AUC = 0.98 for AHP and AUC = 0.96 for LD, but are too complex to allow convenient inspection of their decision-logic.

An interpretable model, denoted M_IDT_, is then obtained by fitting a CART decision tree to *predictions* of the learned full model M_F_ (see e.g. [17] for background on CART models).

Figure 2 displays two decision tree models for LD patient-finding generated using Algorithm IM with two different choices of hyperparameters [38,17]. Each model is accurate, realizing out-of-sample AUC > 0.9 on the training set. Moreover, the models are interpretable: their prediction-logic is straightforward to understand and they offer insight into the way LD presents in GP EHRs. For example, it is seen that LD patients are characterized by high blood glucose and triglycerides (TRI), as expected, and tend to have comorbidities (e.g. diabetes) that are unusual for individuals with normal/low BMI. Perhaps more surprising, presence of ‘clinical confusion’ in a patient’s record, quantified using counts of unique diagnostic codes or consultations, is found to be a useful predictor of LD.

**Figure 2.**
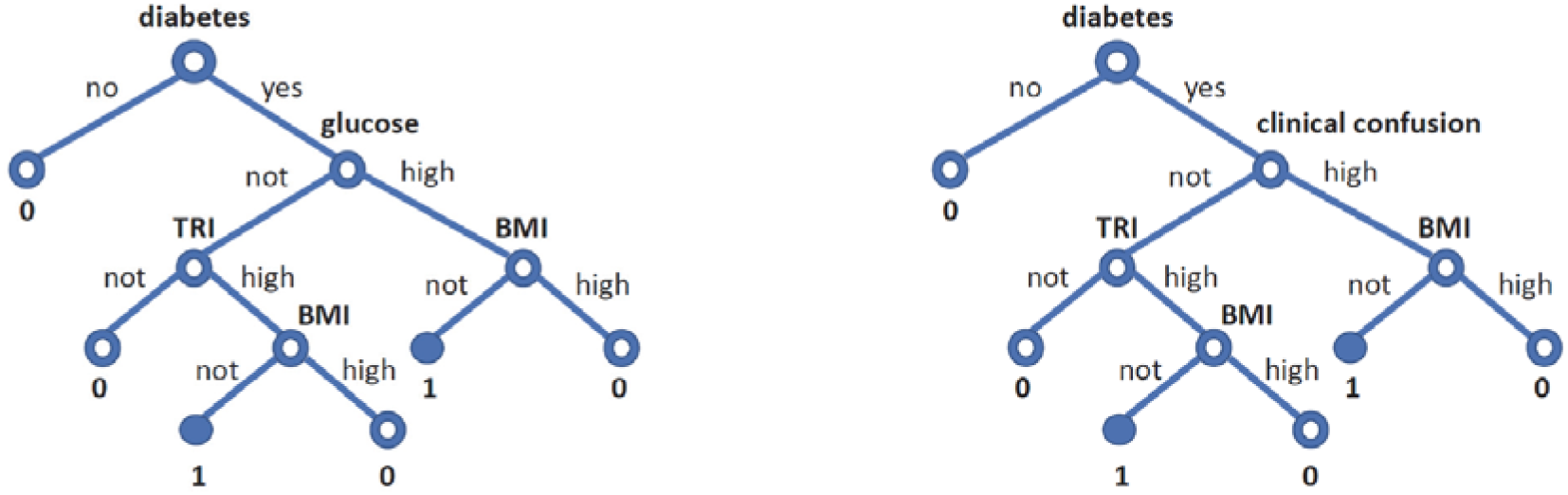
Sample interpretable models for LD patient-finding.

These results broadly agree with, and appear to supplement and increase the utility of, published guidelines for diagnosing LD in clinical settings. For instance, standard guidelines often *begin* with ‘suspicion of LD’ [e.g. 31], which reduces their value to GPs (e.g. there are over 7000 rare diseases, so it may be impractical to require that a GP suspect one of them in order to start the diagnostic process). Papers which may help clinicians ‘suspect LD’ [e.g. 44] have some decision points in common with our models M_IDT_ (e.g. diabetes with normal BMI, high triglycerides) but also miss informative predictors (clinical confusion) and do not specify the decision-logic necessary for implementation.

Because AHP has a heterogeneous clinical presentation [29,30], we built an ensemble of specialist trees to form the interpretable model M_IDT_ for this disease. As an illustrative example, Figure 3 depicts a three-tree ensemble induced by partitioning the full feature space into different subspaces and learning ensemble members on the individual subspaces [38]. It can be seen the individual models either align with medical specialties, such as psychiatry, or reflect features determined to be predictive via the learning process (e.g. feature subspaces associated with a GP examination).

**Figure 3.**
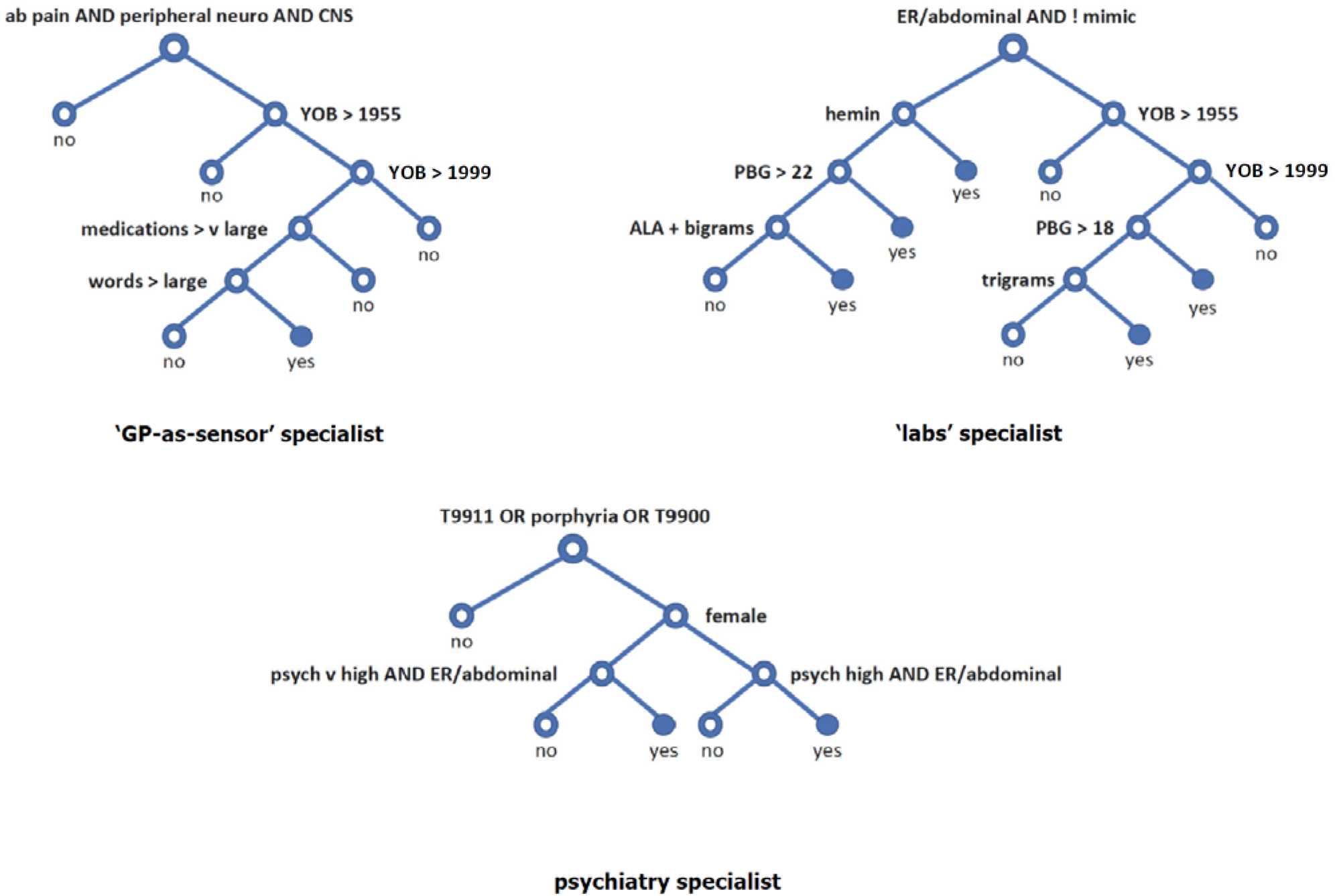
Sample ensemble of interpretable models for AHP patient-finding.

In particular, the tree at top-left of Figure 3 forms predictions by combining well-known diagnostic and demographic information (e.g. abdominal pain, peripheral neuropathy, year of birth) with proxies for clinical confusion (such lengthy clinical notes, numerous disparate medication prescriptions), and thereby encodes ‘GP as an imperfect sensor’ data. The other trees serve to bring ‘labs data’, ‘psychiatric interactions’, and suspicion of porphyria (captured in ICPC codes) into the patient-finding process. Taken together, the ensemble of trees is accurate, providing out-of-sample AUC > 0.9 on the training set, yet interpretable: clinicians can understand each specialist decision trees and the way in which the trees ‘vote’ to make a prediction regarding AHP status.

## 5. Concluding Remarks

This paper presents a novel lightly-supervised approach to learning models with which to detect rare disease patients in EHR databases. The proposed algorithm leverages unlabeled and unreliably-labeled EHR data to induce accurate models from limited numbers of ‘clean’ labels, thereby addressing a key challenge with rare disease patient-finding. We prove the algorithm is *safe:* adding unlabeled/unreliably-labeled data to the learning procedure produces models which are usually more accurate, and never less accurate, than models learned from the reliably-labeled data alone. Moreover, the method is shown to substantially outperform state-of-the-art models in ‘real-world’ patient-finding experiments involving two different rare diseases and a database of 3.2M GP EHRs. The high-performance models generated through light supervision can be transformed into simpler models that are simultaneously accurate and easily-understandable by non-experts.

## Data Availability

The electronic health record data used in this work is protected by Dutch Privacy Regulations, and cannot be made publically available.

## Notes

### Competing Interest Statement

The authors are shareholders in Volv Global.

### Funding Statement

Volv Global provided support for the research and studies reported in the submitted manuscript.

### Author Declarations

The use of the PHARMO data is controlled by the independent Compliance Committee STIZON/PHARMO Institute. All decisions of the Compliance Committee STIZON/PHARMO Institute are based on the applicable legislation in the Netherlands, e.g. the Personal Data Protection Act and the Medical Treatment Contract Act. Within this legal framework, the Code of Conduct 'Use of Data in Health Research' is an important document for the interpretation of the use of this kind of data for scientific research in the Netherlands, and is approved by the Dutch Data Protection Authority.

